# Age-related Reference Data for Cortical and Trabecular 3D-DXA Parameters: the SEIOMM-3D-DXA Project

**DOI:** 10.64898/2026.03.20.26348873

**Authors:** E Casado, S Di Gregorio, C Valero, J González-Macías, JM Olmos, RM Arboiro-Pinel, M Diaz-Curiel, MA Vázquez-Gámez, M Giner, MJ Montoya-García, M Cortés-Berdonces, E Jodar, M Barceló-Bru, JL Pérez-Castrillón, B García-Fontana, M Muñoz-Torres, P Aguado-Acín, C Tornero, M Sosa, F Hawkins, G Martínez Diaz-Guerra, J Del Pino-Montes, J Malouf, L Humbert, M Bracco, L Del Rio

## Abstract

**Purpose:** Osteoporosis and associated hip fractures are a major public health concern. Dual-energy X-ray Absorptiometry (DXA) remains the diagnostic gold standard, but its areal (a) bone mineral density (BMD) measurements have limited sensitivity, as many fractures occur at T-scores above −2.5. Three-dimensional (3D) DXA provides compartment-specific volumetric parameters of the hip, potentially improving osteoporosis management. This study aimed to establish reference data for 3D-DXA parameters to improve osteoporosis management and investigate potential compartmental imbalances at the hip.

**Method:** This multicenter, cross-sectional, population-based observational study (SEIOMM-3D-DXA project), supported by the Spanish Society for Bone and Mineral Metabolism (SEIOMM), analyzed hip DXA scans from 1366 Spanish men and women across six centers. 3D-DXA analyses were conducted using the 3D-Shaper software (3D-Shaper Medical, Barcelona, Spain), producing estimates of trabecular volumetric (v) BMD and cortical surface (s) BMD. Age- and sex-specific reference curves were generated using the LMS method, and thresholds were established based on regression with T-score values. Moreover, trabecular vBMD and cortical sBMD Z-scores were calculated to evaluate potential compartmental imbalances.

**Results:** The derived aBMD curves closely aligned with the NHANES III Caucasian reference. Sex-specific thresholds for trabecular vBMD and cortical sBMD were established for patient stratification. Z-score comparisons revealed significant discrepancies between trabecular and cortical compartments in 52.0% of females and 48.7% of males, underlining the importance of compartment-specific bone assessment.

**Conclusions:** This study establishes reference curves for clinical interpretation of 3D-DXA parameters and demonstrates the potential of 3D-DXA to capture compartmental imbalances at the hip.

**Mini Abstract:** In this study, hip scans from over a thousand men and women in Spain were analyzed to create normative reference values for 3D-DXA parameters. These results can help doctors better stratify people based on the status of each part of the bone and improve the management of osteoporosis.

## Introduction

Osteoporosis is defined as a systemic skeletal disease characterized by low bone mass and microarchitectural deterioration of bone tissue, with a consequent increase in bone fragility and susceptibility to fracture [1]. Millions of fragility fractures occur worldwide each year, causing significant increases in morbidity and mortality [2]. The number of global deaths attributable to low bone mineral density increased by 111% between 1990 and 2019, indicating that the societal burden of osteoporosis and the associated bone loss is on the rise [[3] Hip fracture, in particular, is associated with the highest mortality, with a six-fold increase in risk of death in the first six months after the event [4]. Therefore, timely diagnosis and medical management of osteoporosis are paramount.

Clinical guidelines recommend Dual-energy X-ray Absorptiometry (DXA)-derived areal bone mineral density (aBMD) as the clinical gold standard for osteoporosis diagnosis and treatment monitoring [5,6]. For clinical decision making, aBMD is converted to T-score, a standardized number that compares the measured aBMD with young adult reference values. The International Society of Clinical Densitometry (ISCD) recommends using a uniform Caucasian female normative database [7]. For hip DXA, the NHANES III data serves as reference standard in many countries [8,9]. Conventionally, a T-score between −1 and −2.5 indicates low bone density, or osteopenia, and a T-score equal to or less than −2.5 indicates osteoporosis. The T-score calculation is DXA manufacturer-specific.

Therefore, hip DXA evaluation benefits from a wide standardization, which makes aBMD a reliable and simple marker of bone status. However, fracture prediction with aBMD alone lacks sensitivity, because most fractures happen at T-scores above −2.5 [1]. One of the main drawbacks of DXA examination is that it only provides a 2D projection of bone density, biased by bone thickness in the direction perpendicular to the projected area [10] Therefore, 2D DXA may report different aBMD values for bones that have the same true volumetric BMD but different volumes. Moreover, DXA lacks the ability to distinguish between trabecular and cortical bone.

To address these limitations, 3D assessments are needed. Quantitative computed tomography (QCT) provides a 3D reconstruction of bone density, but its use in clinical practice is limited due to high radiation exposure and lack of standardization [11].

Alternative approaches are based on 3D modeling of patient-specific bone geometry and volumetric BMD starting from a standard 2D DXA acquisition of the hip [12,13]. The 3D-DXA algorithm implemented in the 3D-Shaper® software (3D-Shaper Medical, Barcelona, Spain) uses a QCT-based statistical shape and density model that is automatically registered to the patient’s hip DXA scan, providing compartment-specific volumetric parameters [12]. The accuracy of 3D-DXA cortical and trabecular parameters was validated against QCT [12,14,15]. The precision of 3D-DXA parameters was found to be comparable to that of aBMD [16].

Population-based prospective studies revealed significant association between 3D-DXA parameters and fracture risk [17]. In addition, 3D-DXA parameters were shown to capture compartment-specific bone changes in patients receiving osteoporosis drug treatment within randomized control trials or real-world settings [18,19].

Age- and sex-specific reference curves are necessary for clinical interpretation of 3D-DXA parameters. In this study, we collected a large database of DXA scans in the context of the multicenter SEIOMM-3D-DXA project, supported by the Spanish Society for Bone and Mineral Research (in Spanish, Sociedad Española de Investigación Ósea y del Metabolismo Mineral, SEIOMM). 3D-DXA analyses were performed using 3D-Shaper software. Sex- and manufacturer-specific normative data for the 3D-Shaper cortical and trabecular parameters, as well as clinical thresholds equivalent to −1 and −2.5 total hip aBMD T-scores were calculated. Finally, we evaluated the performance of 3D-Shaper in identifying clinically significant discrepancies between cortical and trabecular parameters within the SEIOMM-3D-DXA cohort.

## Methods

### Study subjects

The data for this analysis were collected from six centers^*^ in Spain that participated in the Spanish multicenter, population-based SEIOMM-3D-DXA project (2018-2021), supported by SEIOMM. Study participants were adult men and women aged 20 years or older. The exclusion criteria were the use of osteoporosis medication, the presence of metabolic or chronic diseases that influence bone metabolism (such as rickets, hyperparathyroidism and hypoparathyroidism), a history of fragility fractures, early or surgical menopause, hypogonadism, severe scoliosis and medical history of spine surgery (orthopedic implant, laminectomy, or vertebral augmentation procedures). The study protocol received ethical approval in agreement with the protocol of Helsinki from the ethical committees of the centers participating in the project and informed consent was obtained from all study participants.

### DXA and 3D-DXA analyses

All subjects had a hip DXA scan acquired with iDXA or Prodigy scanners (GE Healthcare, Chicago, USA) according to the recommendation of the manufacturer. Total hip aBMD was calculated using enCORE software (v18, GE Healthcare). T-score was calculated using the U.S. NHANES III reference data for Caucasian population [7].

The hip scans were analyzed using the 3D-Shaper Software (v2.14, 3D-Shaper Medical, Barcelona, Spain) to obtain 3D bone parameters. Briefly, the software performs the fitting of a 3D statistical shape and density model of the proximal femur to the DXA image. The statistical model was built using 3D models derived from QCT scans [12]. The 3D-Shaper cortical and trabecular parameters include trabecular volumetric (v) BMD [mg/cm^3^] and cortical surface (s) BMD [mg/cm^2^], both calculated at the total hip region of interest. Cortical sBMD is calculated as the multiplication of the cortical vBMD [mg/cm^3^] and the cortical thickness [cm].

### Age-related reference curves

Age-related reference curves for trabecular vBMD and cortical sBMD were obtained using the LMS fitting method proposed by Cole and Green (GAMLSS package v5.0, R software v3.3.2) [24]. The fitting model assumes a skew normal distribution of age-dependent measurements, which becomes symmetric by applying Box-Cox power transformations within each age group. Three age-dependent functions (L, M, S, respectively representing data skewness, median and coefficient of variation) are used to fit the reference data and smoothed along with the age groups to avoid unphysiological discontinuities. The obtained mean ± standard deviation curve across the age span considered (20 to 90 years old) is used as reference curve for the considered population. The process was performed separately for male and female subjects, to obtain sex-specific reference curves. Reference curves were also created for the total hip aBMD data collected within this study.

In addition, SEIOMM-3D-DXA total hip aBMD values were compared to NHANES III reference data for Caucasian adults (men and women) using manufacturer-provided reference curves.

### T-score-equivalent thresholds

A regression analysis between 3D-Shaper parameters and total hip aBMD T-scores was performed in order to obtain thresholds for the 3D-Shaper parameters. Sex-specific thresholds corresponding to total hip aBMD T-scores of −1 and −2.5 were calculated for both trabecular vBMD and cortical sBMD. The thresholds allow to characterize the trabecular vBMD and cortical sBMD as normal, low, or very low.

### Z-score difference

The Z-score is an age-dependent index calculated as the distance of the measurement from the mean value at the corresponding age, divided by the reference standard deviation. Trabecular vBMD and cortical sBMD Z-scores for the SEIOMM-3D-DXA subjects were calculated. The difference between the trabecular vBMD and cortical sBMD Z-scores was used to assess possible discrepancy between the two compartments.

To assess the clinical significance of these differences, we evaluated the precision of Z-score difference. Precision estimates were derived from a previous study, in which 30 subjects were scanned twice using an iDXA device (GE Healthcare) [16]. For the present study, these DXA scans were reanalyzed using the same version of the 3D-Shaper software applied in our current analyses. Z-score differences between trabecular vBMD and cortical sBMD were then calculated using the reference data presented here. The least significant change (LSC) for the Z-score difference was calculated as 2.77 times the precision error, which was defined as the root mean square standard deviation of differences between duplicate measurements divided by √2.

### Manufacturer-specific reference data

Because the DXA scans from the SEIOMM-3D-DXA study were acquired using GE Healthcare devices, the reference data calculated for aBMD, trabecular vBMD, and cortical sBMD provide normative values specific to GE Healthcare scanners. To account for the measurement differences between the two main DXA producers, GE Healthcare and Hologic, we performed a conversion of the GE Healthcare total hip aBMD data (g/cm^2^) using the following equation from the literature [25]:

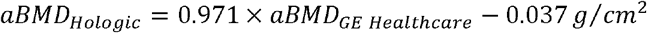

The conversion equations to transform the GE Healthcare 3D-Shaper cortical sBMD and trabecular vBMD measurements into Hologic-equivalent measurements were derived from data of 120 men and women (aged 22-75 years) who underwent scans on both an iDXA (GE Healthcare) and a Discovery (Hologic) scanner at two medical centers, CETIR ASCIRES and Hospital de la Santa Creu i Sant Pau (both in Barcelona, Spain). Linear regression analyses were used to establish the conversion formulas, yielding the following equations:

## Results

### Study subjects

1366 subjects (1015 female and 351 male) were included in the current study. At least 30 subjects per decade were included, except for men of 80 years or older (n=20). **Error! Reference source not found**. in Supplementary Material reports the number of included subjects per sex (F/M) and age groups.

### Age-related reference curves

In **Figure 1** we report the individual measurements and reference curves for total hip aBMD in men and women for the SEIOMM-3D-DXA database (current study). Mean values decreased progressively with age, with a steeper decline after menopause in women. The reference curves from the NHANES III normative database are plotted on top of the current study curves for visual comparison.

**Figure 1.**
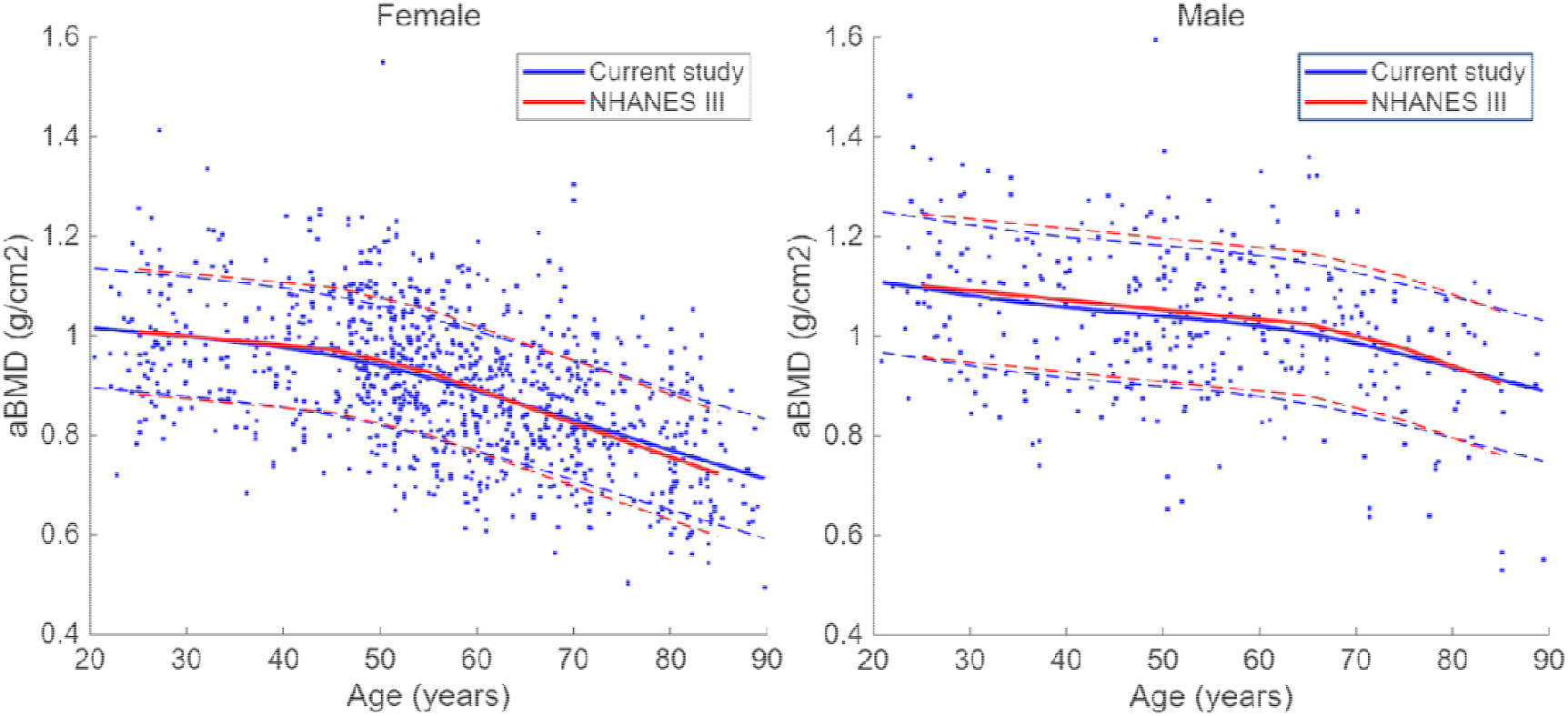
Comparison between SEIOMM-3D-DXA and NHANES III reference curves for Caucasian population. Total hip areal bone mineral density (aBMD, g/) individual values from the female (left) and male (right) cohorts from SEIOMM-3D-DXA (blue dots) are plotted against age, together with the corresponding fitted mean curve (blue, solid) ± SD (dashed), and the corresponding NHANES III reference curve (red, solid) ± SD (dashed).

### T-score-equivalent thresholds

Regression analyses revealed strong correlations (R^2^ =0.88) with aBMD T-score for both cortical sBMD and trabecular vBMD in both sexes (**Figure *2***). Sex-specific thresholds equivalent to aBMD T-scores of −1.0 and −2.5 are reported in Table. These thresholds delineate normal, low, and very low cortical and trabecular density.

**Figure 2.**
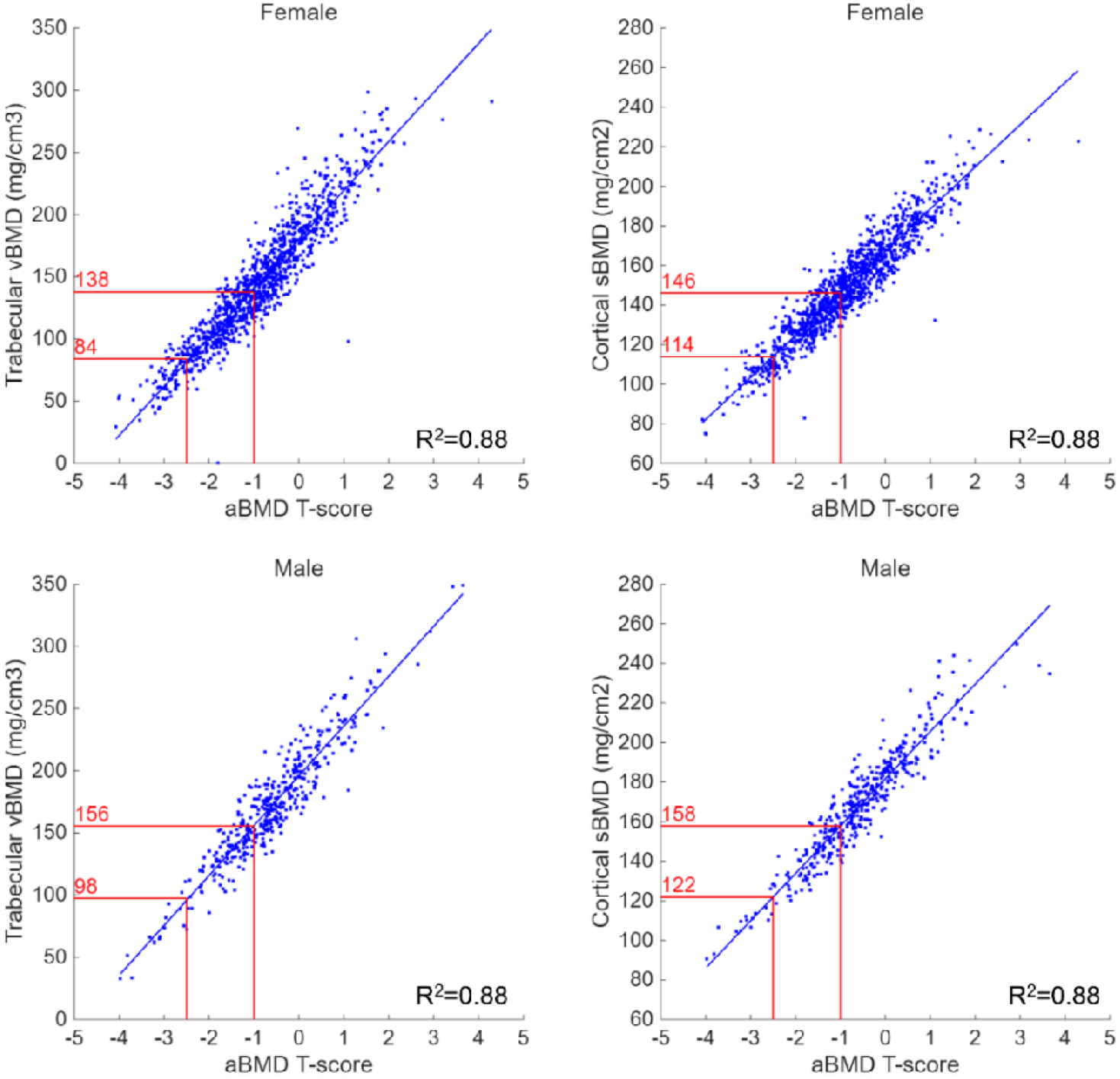
Regression analysis between trabecular vBMD (mg/cm^3^) and aBMD T-score (left) and between cortical sBMD (mg/cm^2^) and aBMD T-score (right), for female (top) and male population (bottom). The threshold values (in red) correspond to aBMD T-score of −1 and −2.5. For all parameters and sexes, the correlation was significant (R^2^ =0.88, p<0.001).

**Table 1.** Sex-specific T-score-equivalent 3D-DXA parameters thresholds.

|  | T-score-equivalent threshold | F | M |
| --- | --- | --- | --- |
| Cortical sBMD (mg/cm<sup>2</sup>) | Normal/low (-1) | 146 | 158 |
|  | Low/very low (-2.5) | 114 | 122 |
| Trabecular vBMD (mg/cm<sup>3</sup>) | Normal/low (-1) | 138 | 156 |
|  | Low/very low (-2.5) | 84 | 98 |

Building up from the results from the regression analysis, a visual representation of the reference curve is provided in **Figure *3***. Green, yellow and red areas on the plot indicate normal, low and very low cortical or trabecular density zones respectively. The black lines are the age-related reference curves (mean ± SD).

**Figure 3.**
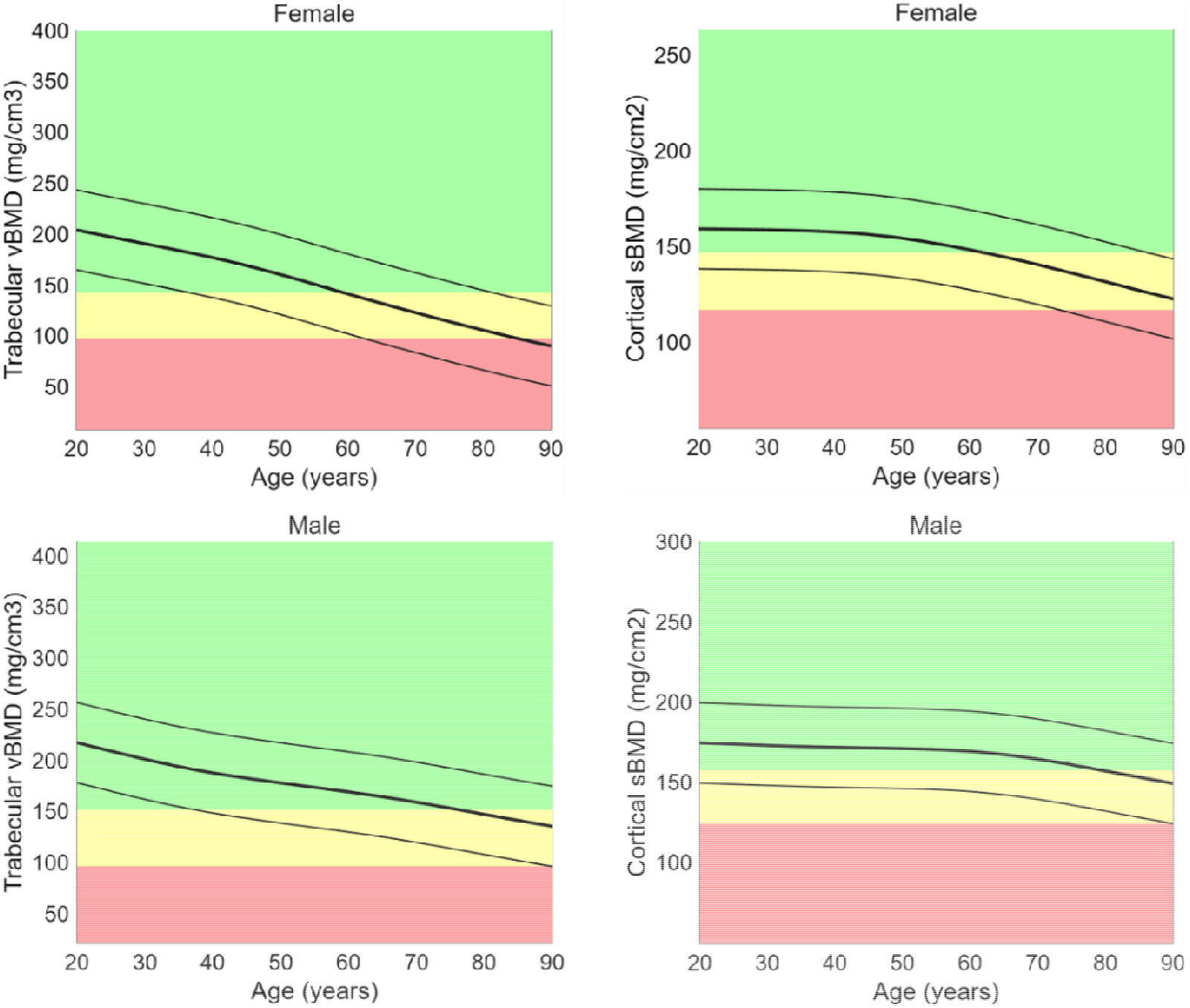
Age-related reference curves for trabecular vBMD (left) (mg/cm^3^), and cortical sBMD (right) (mg/cm^2^); females (top) and males (bottom). The age-related reference data are reported as solid curves. The thicker line represents the mean value at each age, the thin lines are at a distance of ± one standard deviation from the mean curve. The green, yellow, and red regions correspond to normal, low, and very low cortical sBMD or trabecular vBMD, respectively.

In the female population, cortical sBMD starts decreasing after the age of 40 and continues decreasing steadily, with an average loss of −0.94 mg/cm^2^ per year after 65 years old (**Error! Reference source not found**. in Supplementary Material). In the male population, the decrease is found to start later, at about 60 years, and is less pronounced, with an average loss of −0.74 mg/cm^2^ per year after 65 years old. The loss of trabecular vBMD is also more pronounced in women, with a steady decrease throughout adult life, getting steeper around 50 years of age, with average loss of −1.73 mg/cm^3^ per year after 65 years old, compared to −1.21 mg/cm^3^ per year in male subjects.

A comparison of the reference curves calculated for female and male subjects is presented in **Figure *4***. The reference curves from aBMD and the two 3D-DXA parameters for the two sexes are displayed as overlay plots in green (males) and red (females) with the respective standard deviations.

**Figure 4.**
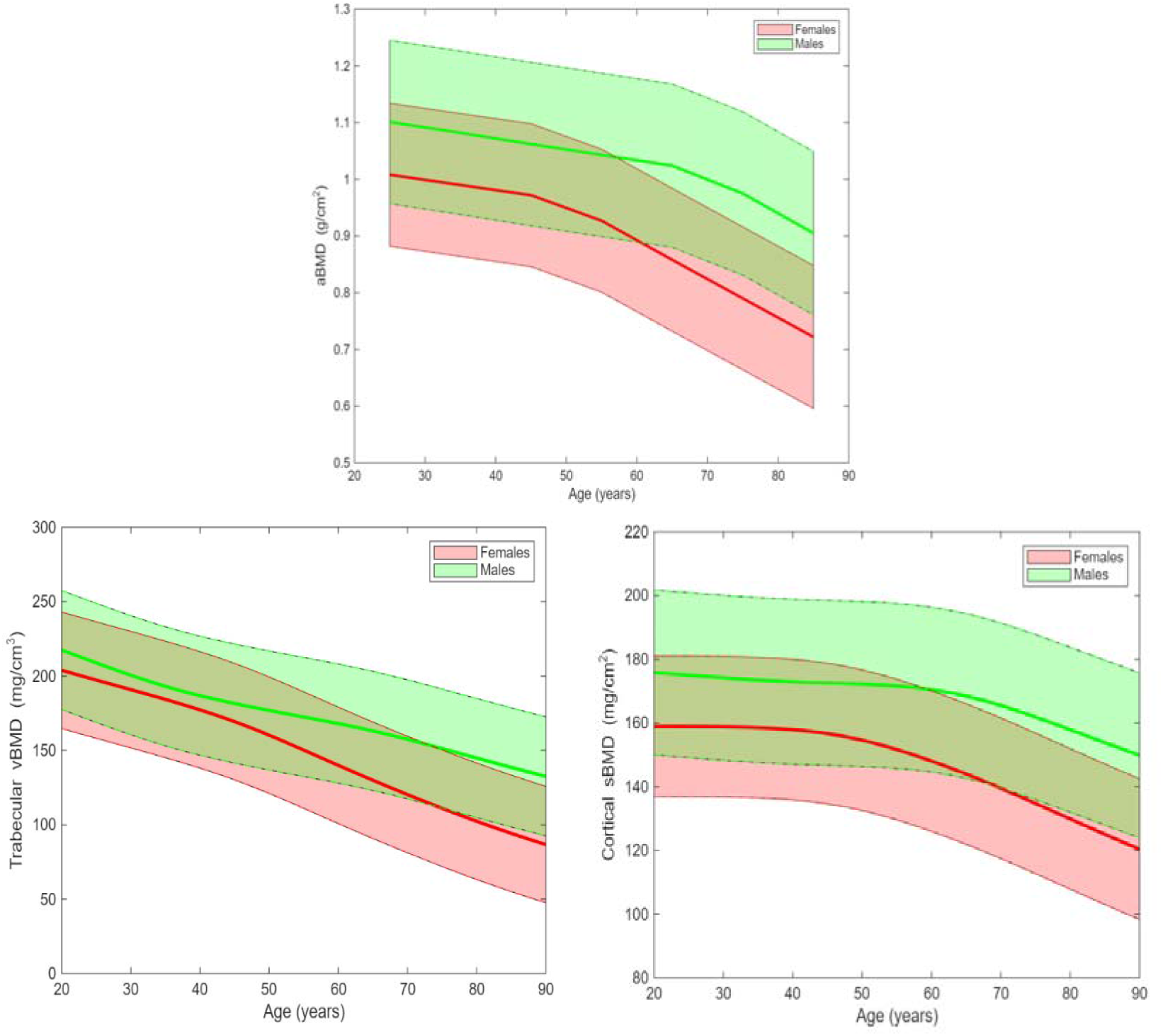
Comparison between reference curves for female (red) and male (green) subjects for aBMD (top), trabecular vBMD (bottom left) (mg/cm^3^), and cortical sBMD (bottom right) (mg/cm^2^). The shaded area is delimited by mean ± standard deviation of the reference curves.

### Z-score differences

The histograms in **Figure *5*** show the distribution of the differences between cortical sBMD and trabecular vBMD Z-scores, for females and males. The LSC for the Z-score difference was 0.397, as calculated using data from a previous precision study [16]. 52.0% of females and 48.7% of males showed an absolute Z-score difference larger than the LSC. Specifically, 26.3% of females and 25.4% of males (red bins in **Figure *5***) had higher cortical sBMD Z-scores, while the rest (25.7% of females and 23.4% of males, green bins in **Figure *5***) had higher trabecular vBMD Z-scores.

**Figure 5.**
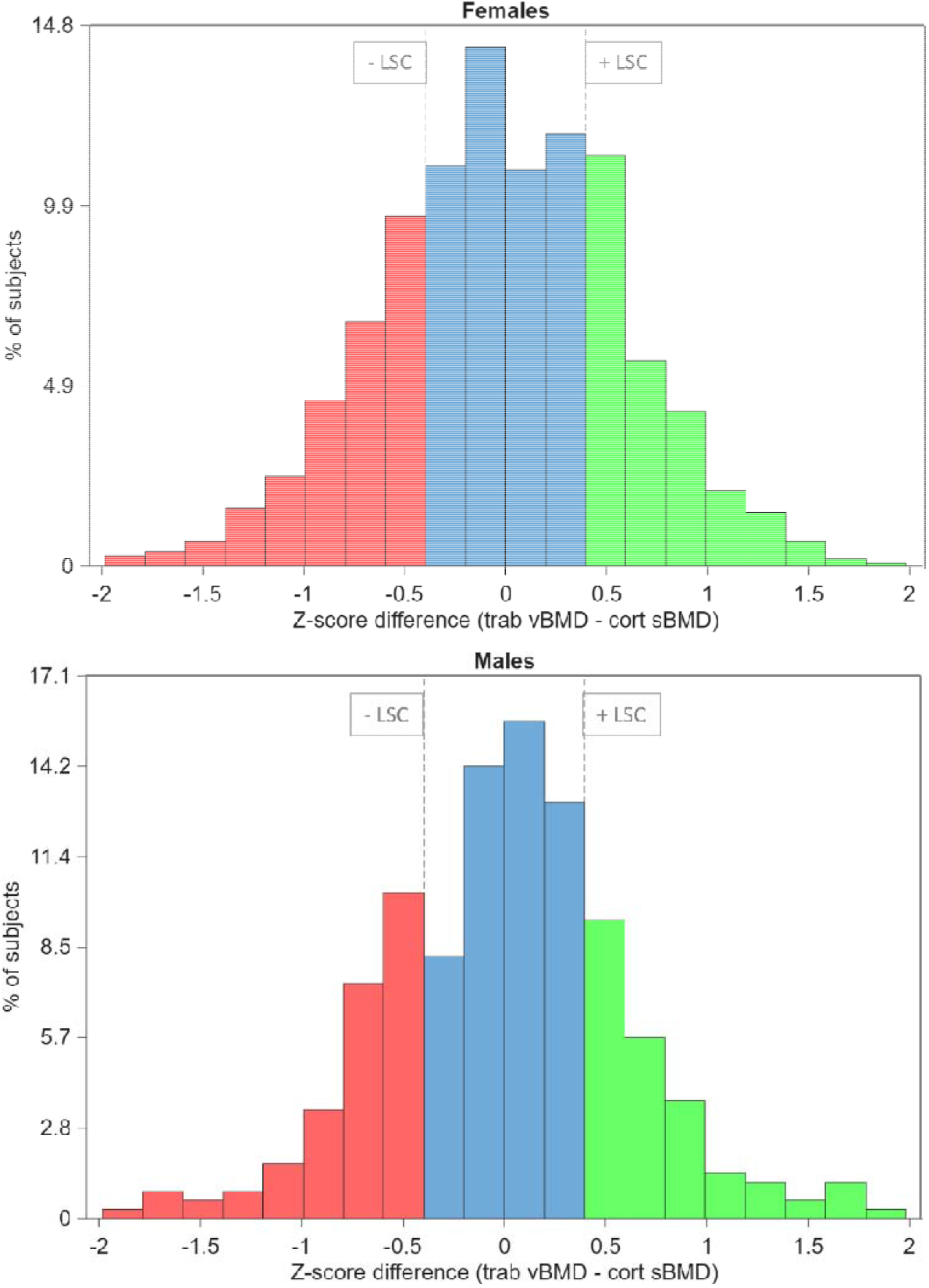
Z-score differences distribution in female (top) and male (bottom) subjects, expressed as percentage of the total (1011 females and 351 males). The least significant change (LSC) for the Z-score difference was calculated from a previous precision study [16]. + LSC and – LSC are represented as two vertical dashed lines. Z-score differences higher than + LSC (green) indicate a clinically significant higher trabecular vBMD Z-score, while Z-score differences lower than - LSC (red) indicate a significantly higher cortical sBMD Z-score.

### Conversion to Hologic data

The reference data obtained after applying the conversion from GE Healthcare to Hologic data are reported in the Supplementary Material Figures S1, S2, S3 and Table S3.

## Discussion

The multicenter SEIOMM-3D-DXA project, initiated in 2018, is the first study aiming at establishing reference data for the 3D-Shaper trabecular and cortical parameters. The results support clinical interpretation of 3D-Shaper parameters, indicating how individual compartment-specific measurements compare with reference data. In addition, to align with current standard clinical practice, thresholds analog to aBMD T-scores are presented. To the best of our knowledge, this is the first study proposing clinical thresholds for the 3D-Shaper measurements.

The presented methodology employed the LMS method for determining sex-specific age-related curves. This approach is consistent with previous works on other bone indices such as trabecular bone score (TBS) and high-resolution peripheral QCT (HR-pQCT) parameters [26–30]. However, unlike TBS, the strong correlation between 3D-DXA parameters and aBMD allowed thresholds to be established through regression analyses between 3D-DXA parameters and aBMD T-scores.

In general, men had a higher overall hip aBMD, and females experienced steeper decrease in later life, after menopause. Similarly, a more pronounced decrease in cortical sBMD existed in female subjects, with an earlier onset (around 50 years old) compared to male subjects (around 60 years old). Conversely, trabecular vBMD was found to decrease steadily in subjects aged 20 years or older, i.e. after reaching peak bone mass. This was the case for both male and female subjects, although the total decrease was higher in females. These tendencies are in line with a previous multicenter study on 1354 healthy adults, endorsed by the Argentinian Society of Osteology and Mineral Metabolism (AAOMM), aimed at determining reference curves for trabecular vBMD and cortical sBMD for the Argentinian population [31].

Another study on 2647 Spanish people (65% females) recruited from a large single-center cohort (Camargo Cohort, Santander, Spain) found no significant differences in trabecular vBMD in the two sexes before the age of 60, while DXA aBMD differs [32]. The same trends were observed in our analysis. Trabecular vBMD curves were very similar between the sexes in subjects under 50 years of age, whereas women showed lower total hip aBMD compared to men (Figure 4). DXA-based aBMD is influenced by bone size, with larger bones producing higher aBMD values. This may explain the sex-related differences in aBMD observed both in our study and in the Camargo Cohort study. The similar trabecular vBMD curves in men and women further support that 3D-DXA trabecular vBMD provides a true volumetric density measurement, independent of bone size.^32^ After the age of 65 years, yearly trabecular vBMD loss became steeper in women, while remaining stable in men.

Age-related bone loss in the Japanese population was also investigated using 3D-Shaper [33]. This study including 1372 elderly Japanese subjects revealed a decrease of 8.4% in trabecular vBMD and 9.6% in cortical sBMD in 65 to 80 years old women, with an accentuated loss in both parameters after the age of 70. In elderly men, no significant differences were found across age groups, although in both parameters the cumulated loss was larger than 5%. Our study revealed comparable trends, with male subjects exhibiting a slower decline in both compartments compared to females after the age of 65.

Another important finding of our study is that the aBMD reference curves derived from participants in the SEIOMM-3D-DXA project closely matched the NHANES III curves for Caucasian adults. A similar observation was previously reported in the OsteoSER project, a screening program conducted in the Spanish population [34]. This finding demonstrates that our normative data are both representative of the Spanish population and consistent with the standard NHANES III reference data, which the ISCD recommends for T-score calculation and interpretation of hip DXA measurements. These results also support the combined use of NHANES III data for aBMD alongside SEIOMM-2D-DXA reference curves for 3D-DXA cortical and trabecular parameters interpretation.

Importantly, the SEIOMM-3D-DXA age-related reference data and thresholds presented in the current study are integrated in the 3D-Shaper software version 2.14. This software version has received regulatory clearance in multiple regions, including the U.S., Europe, Asia (Japan, Singapore, and Thailand), and LATAM (Brazil and Argentina), supporting its clinical use in osteoporosis management.

The influence of the race or ethnic group on BMD and fracture risk is still an open debate in the scientific community. Although ISCD recommends the use of U.S. data from NHANES III as the reference standard for DXA-based measurements at femoral neck and total hip, regardless of the ethnic group, it has been shown that Asian populations have a lower BMD and bone size, and Black population tend to have higher BMD [7,35,36]. In a previous study with 3D-Shaper, Black women had significantly higher trabecular vBMD and cortical sBMD than White women, despite being older [37]. Further research is warranted to determine whether race- or ethnicity-specific normative values are necessary for 3D-DXA parameters.

A previous study reported a strong correlation between 3D-DXA cortical and trabecular parameters and aBMD [38]. In the same study, it was also hypothesized that discrepancies observed between trabecular and cortical parameters in individual subjects may be attributable to measurement precision errors. In the present study, we likewise found a high correlation between 3D-DXA parameters and aBMD. However, analysis of the differences between trabecular and cortical Z-scores revealed that 52.0% of women and 48.7% of men exhibited a clinically significant discrepancy between the two compartments. This finding indicates that a substantial proportion of individuals show differences between cortical and trabecular parameters that are not attributable to measurement precision errors. These significant inter-compartmental imbalances cannot be captured by standard DXA, undermining the interpretation of clinical results, especially in patients with secondary osteoporosis or under treatment. In several previous studies, 3D-DXA parameters were shown to provide meaningful insight in the management of secondary osteoporosis, osteoporotic fracture risk, and treatment monitoring [17–23, 37, 39-43]. Further research is needed to investigate the potential clinical impact of imbalances between cortical and trabecular parameters on bone strength, fracture risk, treatment decision-making, and monitoring.

Some limitations need to be highlighted. First, the SEIOMM-3D-DXA project was conducted in Spain, which limits the generalizability of the present findings to other regions, races, or ethnicities. However, the aBMD SEIOMM-3D-DXA data closely aligned with the NHANES III curves for Caucasian adults, which are recommended by the ISCD for T-score calculation and hip DXA interpretation. Future studies will aim to extend these analyses to diverse populations and ethnicities. Second, our comparison with NHANES III was restricted to aBMD, as 3D-Shaper could not be applied to the older DXA scans from NHANES III due to compatibility limitations. Third, reference curves were calculated from GE Healthcare scanners, while Hologic reference curves were derived through conversion equations. Although no Hologic data were included, the aBMD curves still closely matched NHANES III curves for Caucasian adults across both sexes and scanner manufacturers, as shown in Figures 3 and S3.

In summary, with the present work, we determined the first sex- and manufacturer-specific age-related reference data and thresholds for 3D-DXA parameters, providing a foundation for clinical decision-making based on cortical and trabecular bone status. The observed high prevalence of discrepancies between compartments emphasizes the value of compartment-specific evaluation. Further research should explore race- and ethnicity-specific reference ranges and the clinical relevance of inter-compartmental imbalances.

## Data Availability

The reference curves obtained from this study will be made available upon reasonable request.

## Acknowledgements

authors acknowledge the Spanish Society for Bone and Mineral Metabolism (SEIOMM) for its support of the multi-center SEIOMM-3D-DXA project.

## Ethics approval statement

The study protocol received ethical approval in agreement with the protocol of Helsinki from the ethical committees of the centers participating in the project.

## Patient consent statement

Informed consent was obtained from all study participants.

## Footnotes

* Hospital Universitari Parc Taulí, Sabadell; CETIR Grup Mèdic, Barcelona; Hospital Ruber Juan Bravo Quirón Salud, Madrid; Hospital Universitario Vall d’Hebron, Barcelona; Hospital Rio Hortega, Valladolid; Hospital Universitario La Paz, Madrid.

## References

1. Compston JE, McClung MR, Leslie WD (2019) Osteoporosis. Lancet 393(10175):1019–31. 10.1016/S0140-6736(18)32112-3

2. Sattui SE, Saag KG (2014) Fracture mortality: Associations with epidemiology and osteoporosis treatment. Nat Rev Endocrinol 10:592–602. 10.1038/nrendo.2014.125

3. Shen Y, Huang X, Wu J et al (2022) The Global Burden of Osteoporosis, Low Bone Mass, and Its Related Fracture in 204 Countries and Territories, 1990-2019. Front Endocrinol 13:882241. 10.3389/fendo.2022.882241

4. Abrahamsen B, Van Staa T, Ariely R, Olson M, Cooper C (2009) Excess mortality following hip fracture: A systematic epidemiological review. Osteoporos Int 20(10):1633–50. 10.1007/s00198-009-0920-3

5. Cosman F, de Beur SJ, LeBoff MS et al (2014) Clinician’s Guide to Prevention and Treatment of Osteoporosis. Osteoporos Int 25(10):2359–81. 10.1007/s00198-014-2794-2

6. Compston J, Cooper A, Cooper C et al (2017) UK clinical guideline for the prevention and treatment of osteoporosis. Arch Osteoporos 12(1):43. 10.1007/s11657-017-0324-5

7. Shuhart C, Cheung A, Gill R, Gani L, Goel H, Szalat A (2024) Executive Summary of the 2023 Adult Position Development Conference of the International Society for Clinical Densitometry: DXA Reporting, Follow-up BMD Testing and Trabecular Bone Score Application and Reporting. J Clin Densitom 27(1):101435. 10.1016/j.jocd.2023.101435

8. Kelly TL, Wilson KE, Heymsfield SB (2009) Dual energy X-ray absorptiometry body composition reference values from NHANES. PLoS One 4(9):e7038. 10.1371/journal.pone.0007038

9. Kiebzak GM, Binkley N, Lewiecki EM, Miller PD (2007) Diagnostic Agreement at the Total Hip Using Different DXA Systems and the NHANES III Database. J Clin Densitom 10(2):132–7. 10.1016/j.jocd.2007.02.003

10. Nicks KM, Anderson PA, Libber J et al (2013) Three-dimensional structural analysis of the proximal femur in an age-stratified sample of women. Bone 55(1):179–88. 10.1016/j.bone.2013.02.009

11. Wang L, Yin L, Zhao Y et al (2019) QCT of the femur: Comparison between QCTPro CTXA and MIAF Femur. Bone 120:262–70. 10.1016/j.bone.2018.10.016

12. Humbert L, Martelli Y, Fonolla R et al (2017) 3D-DXA: Assessing the Femoral Shape, the Trabecular Macrostructure and the Cortex in 3D from DXA images. IEEE Trans Med Imaging 36(1):27–39. 10.1109/TMI.2016.2593346

13. Väänänen SP, Grassi L, Flivik G, Jurvelin JS, Isaksson H (2015) Generation of 3D shape, density, cortical thickness and finite element mesh of proximal femur from a DXA image. Med Image Anal 24(1):125–34. 10.1016/j.media.2015.06.001

14. Sone T, Humbert L, Lopez Picazo M, Winzenrieth R, Ohnaru K (2021) Assessment of femoral shape, trabecular and cortical bone in Japanese subjects using DXA-Based 3D modelling. J Bone Miner Res 37 (Suppl 1)

15. Lopez Picazo M, Humbert L, Winzenrieth R, Black D (2022) Accuracy of Femoral Shape, Trabecular and Cortical bone measurements using DXA-based 3D modelling in subjects from the US. J Bone Miner Res 38 (Suppl 1)

16. Humbert L, Winzenrieth R, Guglielmi G et al (2019) 3D Analysis of Cortical and Trabecular Bone From Hip DXA: Precision and Trend Assessment Interval in Postmenopausal Women. J Clin Densitom 22(2):214–8. 10.1016/j.jocd.2018.05.001

17. Iki M, Winzenrieth R, Tamaki J et al (2021) Predictive ability of novel volumetric and geometric indices derived from dual-energy X-ray absorptiometric images of the proximal femur for hip fracture compared with conventional areal bone mineral density: the Japanese Population-based Osteoporosis (JPOS) Cohort Study. Osteoporos Int 32(11):2289–99. 10.1007/s00198-021-06013-2

18. Hadji P, Kamali L, Thomasius F et al (2024) Real-world efficacy of a teriparatide biosimilar (RGB-10) compared with reference teriparatide on bone mineral density, trabecular bone score, and bone parameters assessed using quantitative ultrasound, 3D-SHAPER® and high-resolution peripheral computer tomography in postmenopausal women with osteoporosis and very high fracture risk. Osteoporos Int 35(12):2107–16. 10.1007/s00198-024-07208-z

19. Winzenrieth R, Humbert L, Di Gregorio S et al (2018) Effects of osteoporosis drug treatments on cortical and trabecular bone in the femur using DXA-based 3D modeling. Osteoporos Int 29(10):2323–33. 10.1007/s00198-018-4624-4

20. Cole TJ, Green PJ (1992) Smoothing reference centile curves: The lms method and penalized likelihood. Stat Med 11(10):1305–19. 10.1002/sim.4780111005

21. Binkley N, Kiebzak GM, Lewiecki EM et al (2005) Recalculation of the NHANES database SD improves T-score agreement and reduces osteoporosis prevalence. J Bone Miner Res 20(2):195–201. 10.1359/JBMR.041115

22. Simonelli C, Leib E, Mossman N et al (2014) Creation of an age-adjusted, dual-energy x-ray absorptiometry-derived trabecular bone score curve for the lumbar spine in non-hispanic US white women. J Clin Densitom 17(2):314–9. 10.1016/j.jocd.2013.09.002

23. Dufour R, Winzenrieth R, Heraud A, Hans D, Mehsen N (2013) Generation and validation of a normative, age-specific reference curve for lumbar spine trabecular bone score (TBS) in French women. Osteoporos Int 24(11):2837–46. 10.1007/s00198-013-2384-8

24. Iki M, Tamaki J, Kadowaki E et al (2015) Age-related normative values of trabecular bone score (TBS) for Japanese women: the Japanese Population-based Osteoporosis (JPOS) study. Osteoporos Int 26(1):245–52. 10.1007/s00198-014-2856-5

25. Whittier DE, Burt LA, Hanley DA, Boyd SK (2020) Sex- and Site-Specific Reference Data for Bone Microarchitecture in Adults Measured Using Second-Generation HR-pQCT. J Bone Miner Res 35(11):2151–8. 10.1002/jbmr.4114

26. Warden SJ, Liu Z, Fuchs RK, van Rietbergen B, Moe SM (2022) Reference data and calculators for second-generation HR-pQCT measures of the radius and tibia at anatomically standardized regions in White adults. Osteoporos Int 33(4):791–806. 10.1007/s00198-021-06164-2

27. Brance ML, Brun LR, Lance MET et al (2024) Age- and Sex-Related Volumetric Density Differences in Trabecular and Cortical Bone of the Proximal Femur in Healthy Population. J Bone Metab 31(4):279–89. 10.11005/jbm.24.765

28. Valero C, Winzenrieth R, Casado E et al (2021) 3D analysis of bone mineral density in a cohort: age- and sex-related differences. Arch Osteoporos 16(1):80. 10.1007/s11657-021-00921-w

29. Otsuka H, Kimbara K, Sato M et al (2025) Age-related differences in bone structural parameters using 3D-DXA and TBS in men and women: The Bunkyo Health Study. Bone 199:117549. 10.1016/j.bone.2025.117549

30. Gómez-Vaquero C, Winzenrieth R, Naranjo A et al (2025) An update in bone mineral density status in Spain: the OsteoSER study. Arch Osteoporos 20(1):37. 10.1007/s11657-025-01520-9

31. Kim KM, Choi SH, Lim S et al (2011) Differences in femoral neck geometry associated with age and ethnicity. Osteoporos Int 22(7):2165–74. 10.1007/s00198-010-1459-z

32. Wu Q, Dai J (2023) Racial/Ethnic Differences in Bone Mineral Density for Osteoporosis. Curr Osteoporos Rep 21:397–408. 10.1007/s11914-023-00838-y

33. Jain RK, López Picazo M, Humbert L, Dickens L, Vokes T (2025) Bone Structural Parameters as Measured by 3-Dimensional Dual-Energy X-Ray Absorptiometry Are Superior in Black Women and Demonstrate Unique Associations With Prior Fracture Versus White Women. Endocr Pract 31(2):152–8. 10.1016/j.eprac.2024.10.015

34. Huininga K, Koromani F, Zillikens MC, van Velsen EFS, Rivadeneira F (2025) Use of DXA-derived 3D-modeling, as implemented by 3D-Shaper, for the assessment of fracture risk in a population-based setting. J Bone Miner Res (In Press). 10.1093/jbmr/zjaf120

35. Winzenrieth R, Ominsky MS, Wang Y, Humbert L, Weiss RJ (2021) Differential effects of abaloparatide and teriparatide on hip cortical volumetric BMD by DXA-based 3D modeling. Osteoporos Int 32(3):575–83. 10.1007/s00198-020-05806-1

36. Winzenrieth R, Kostenuik P, Boxberger J, Wang Y, Humbert L (2022) Proximal Femur Responses to Sequential Therapy With Abaloparatide Followed by Alendronate in Postmenopausal Women With Osteoporosis by 3D Modeling of Hip Dual-Energy X-Ray Absorptiometry (DXA). JBMR Plus 6(4):e10612. 10.1002/jbm4.10612

37. Lewiecki EM, Miller PD, Humbert L et al (2025) Effects of romosozumab and teriparatide on hip bone using 3D-SHAPER in postmenopausal women with osteoporosis. JBMR Plus (In Press). 10.1093/jbmrpl/ziaf151

38. Lewiecki EM, Dinavahi RV, Miller PD et al (2024) 3D-modeling from hip DXA shows improved bone structure with romosozumab followed by denosumab or alendronate. J Bone Miner Res 39(4):473–83. 10.1093/jbmr/zjae028

