## Supplementary Material for "Age-related Reference Data for Cortical and Trabecular 3D-DXA Parameters: the SEIOMM-3D-DXA Project"

**Supplemental Material**

**Table S1** Number of subjects included in the analysis per age group and sex.

| Age group (years old) | Females | Males |
| --- | --- | --- |
| [20-29] | 74 | 42 |
| [30-39] | 63 | 45 |
| [40-49] | 163 | 58 |
| [50-59] | 288 | 84 |
| [60-69] | 205 | 58 |
| [70-79] | 133 | 44 |
| >80 | 89 | 20 |
| Total | 1015 | 351 |

.

**Table S2** Sex-specific average yearly changes in cortical sBMD and trabecular vBMD.

|  | Females | Males |
| --- | --- | --- |
| Cortical sBMD (20 to 50 years old) | -0.15 mg/cm^2^ per year | -0.12 mg/cm^2^ per year |
| Cortical sBMD (65 years or older) | -0.94 mg/cm^2^ per year | -0.74 mg/cm^2^ per year |
| Trabecular vBMD (20 to 50 years old) | -1.46 mg/cm^3^ per year | -1.35 mg/cm^3^ per year |
| Trabecular vBMD (65 years or older) | -1.73 mg/cm^3^ per year | -1.21 mg/cm^3^ per year |

**Table S3** Sex-specific T-score-equivalent 3D-shaper thresholds for Hologic data.

|  | T-score-equivalent threshold | F | M |
| --- | --- | --- | --- |
| Cortical sBMD (mg/cm^2^) | Normal/low (-1) | 145 | 155 |
|  | Low/very low (-2.5) | 117 | 124 |
| Trabecular vBMD (mg/cm^3^) | Normal/low (-1) | 156 | 171 |
|  | Low/very low (-2.5) | 109 | 121 |

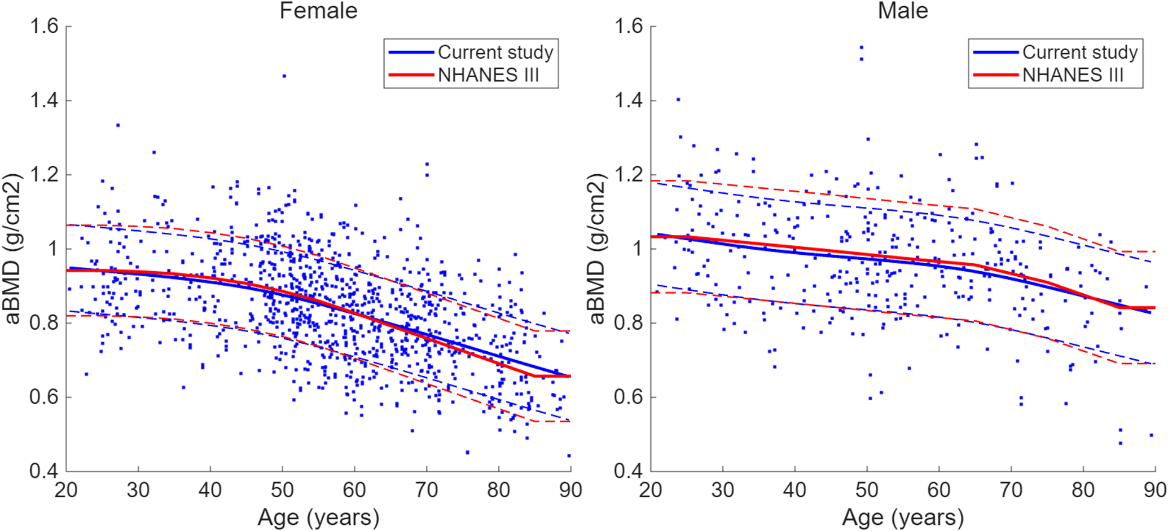

**Figure S1** Comparison between SEIOMM-3D-DXA converted to Hologic-equivalent data and NHANES III Hologic reference curves. Total hip areal bone mineral density (aBMD, g/cm^2^) individual values from the female (left) and male (right) cohorts from SEIOMM-3D-DXA (blue dots) are plotted against age, together with the corresponding fitted mean curve (blue, solid) ± SD (dashed), and the corresponding NHANES III reference curve (red, solid) ± SD (dashed).

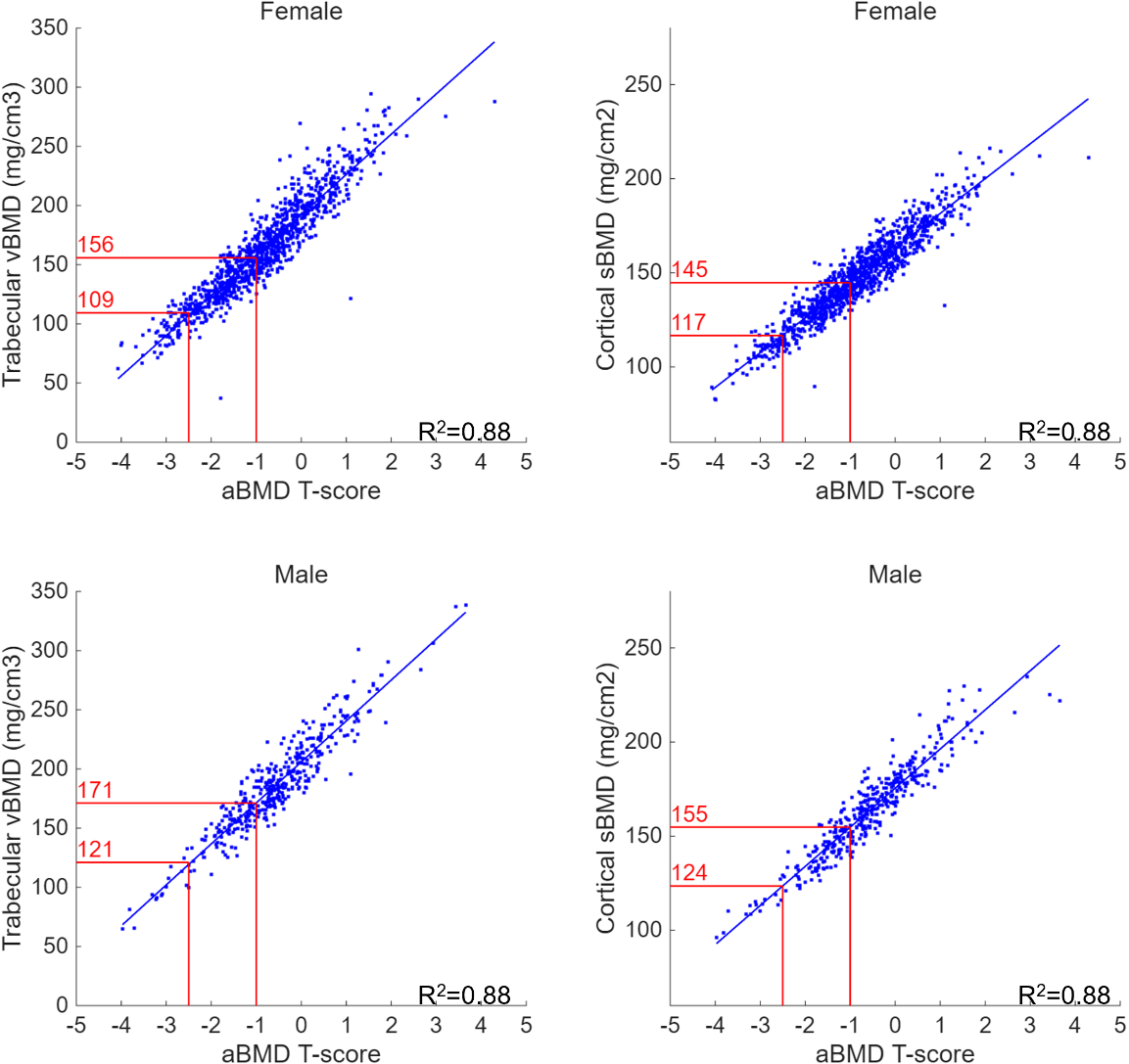

**Figure S2** Regression analysis between cortical sBMD (mg/cm^2^) and aBMD T-score (left) and between trabecular vBMD (mg/cm^3^) and aBMD T-score (right) converted to Hologic-equivalent data, for female (top) and male population (bottom). The threshold values (in red) correspond to aBMD T-score of -1 and -2.5. For all parameters and sexes, the correlation was significant (R^2^=0.88, p<0.001).

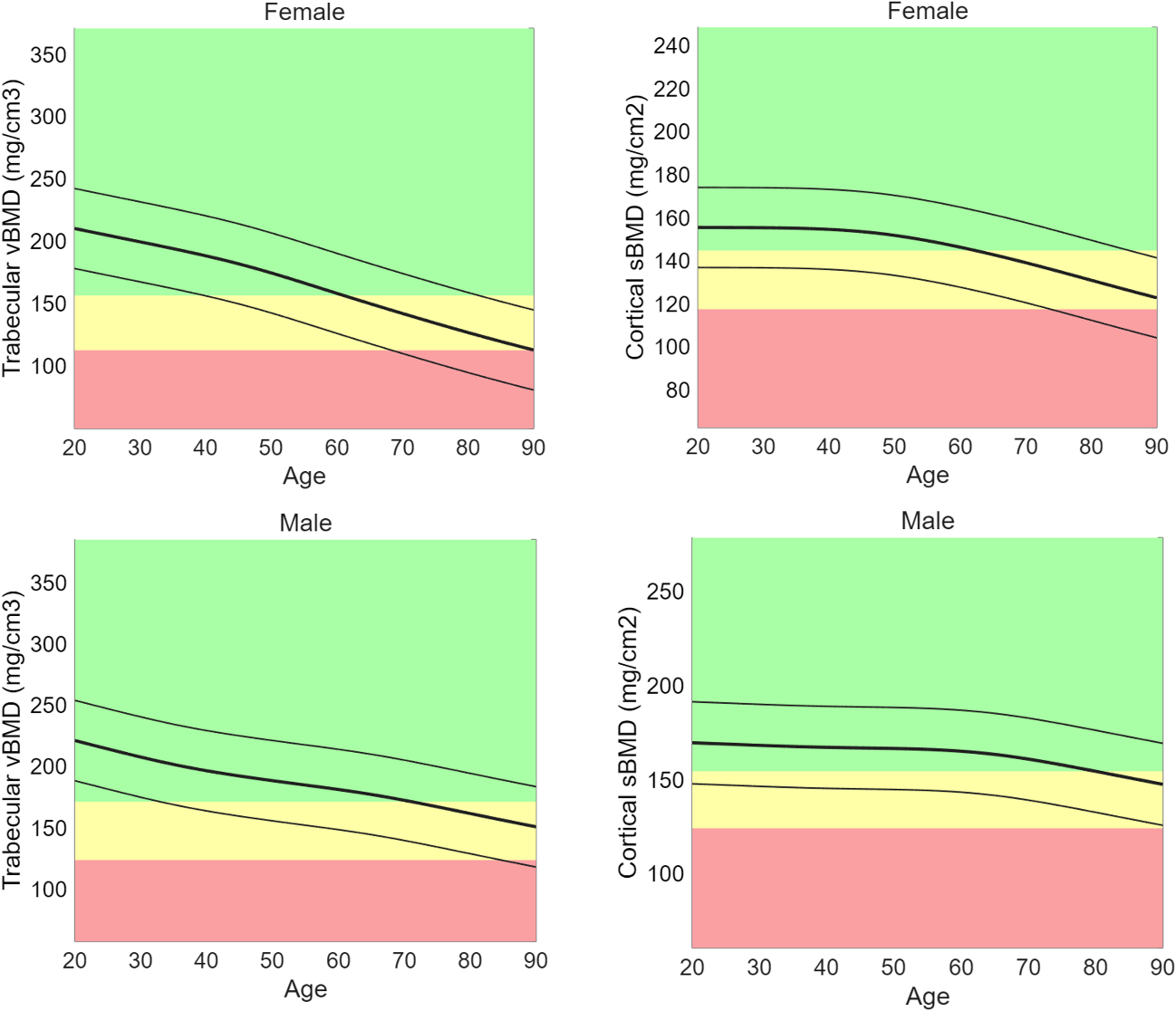

**Figure S3** Age-related reference curves for trabecular vBMD (left) (mg/cm^3^), and cortical sBMD (right) (mg/cm^2^); females (top) and males (bottom) converted to Hologic-equivalent data. The age-related reference data are reported as solid curves. The thicker line represents the mean value at each age, the thin lines are at a distance of ± one standard from the mean curve. The green, yellow, and red regions correspond to normal, low, and very low cortical sBMD or trabecular vBMD, respectively.
